# Food Addiction Symptoms in Adults with Alcohol Use Disorder

**DOI:** 10.64898/2026.08.11.26360166

**Authors:** Jennifer J. Barb, Li Yang, Jenny Yarmovsky, Melanie Schwandt, Vijay Ramchandani, Nancy Diazgranados, Ashley N. Gearhardt, Lorenzo Leggio

## Abstract

**Importance:** Food addiction (FA) has been proposed as a phenotype sharing features with substance use disorders. Despite increasing recognition of food addiction as a behavioral phenotype with features overlapping substance use disorders, little is known about its prevalence or clinical significance among individuals with alcohol use disorder (AUD).

**Objective:** To examine the prevalence of FA and to evaluate demographic, psychological, and alcohol related correlates in individuals with AUD.

**Design, Setting, and Participants:** This cross-sectional analysis included 743 adults with AUD who were either treatment-seeking (Tx) (n = 534) for AUD and were enrolled in an inpatient program at the National Institutes of Health Clinical Center or non–treatment-seeking (non-Tx) (n = 209).

**Main Outcomes and Measures:** FA symptoms were assessed using the Yale Food Addiction Scale, with ≥2 symptoms categorized as FA in this report. Multivariable logistic regression models adjusted for age, education, and income were conducted separately within each cohort.

**Results:** Among 743 adults with AUD, 238 (32.1%) met criteria for FA symptoms, with similar prevalence among Tx (32.6%) and non-Tx (30.6%) participants despite marked differences in clinical characteristics. Across both cohorts, FA was independently associated with higher body mass index, greater psychological distress, and greater alcohol dependence severity. Childhood trauma and poorer sleep quality were additionally associated with FA among treatment-seeking participants, whereas alcohol-related measures differed according to treatment status.

**Conclusions and Relevance:** FA was common among adults with AUD and was associated with greater psychological, behavioral, and metabolic burden regardless of treatment-seeking status. These findings suggest that FA identifies a clinically meaningful subgroup of individuals with AUD who may benefit from more comprehensive assessment and integrated treatment approaches.

**KEY POINTS BOX:** *Question:* Are food addiction (FA) symptoms prevalent among individuals with alcohol use disorder (AUD)?

*Findings:* In this cross-sectional study of 743 adults with AUD, approximately one-third exhibited FA symptoms. Similar prevalence was observed among individuals seeking treatment for AUD and those not seeking treatment for AUD despite differences in clinical characteristics. FA symptoms were associated with higher body mass index, greater psychological distress, and greater alcohol dependence severity, with some associations differing by treatment status.

*Meaning:* FA may identify a clinically meaningful subgroup of individuals with AUD, regardless of treatment-seeking status, characterized by greater psychological and addiction-related burden, supporting further investigation of routine assessment and integrated treatment approaches.

Visual Abstract
Visual Abstract Legend: Visual abstract of experimental design, conceptual model, key findings, and implications. AUD, Alcohol Use Disorder; AUDIT, Alcohol Use Disorders Identification Test; AUDIT-C, Alcohol Use Disorders Identification Test-Concise; BMI, body mass index; Tx, treatment-seeking; non-Tx, non-treatment-seeking; YFAS, Yale Food Addiction Scale.

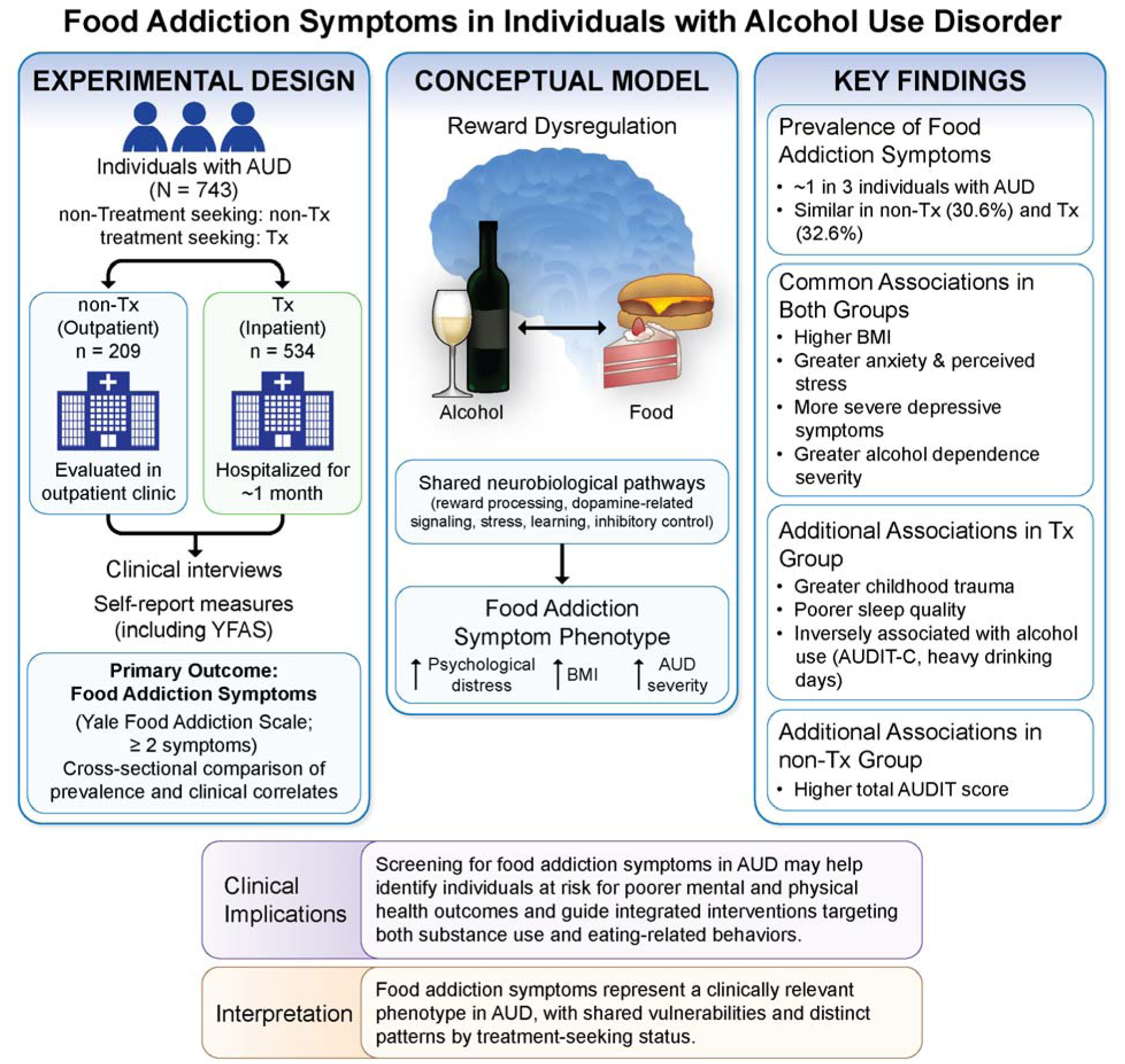

## Introduction

Alcohol use disorder (AUD) affects approximately 27.9 million individuals in the United States and remains a major contributor to preventable morbidity, mortality, and healthcare burden.^1–3^ Similarly, low quality diets characterized by high consumption of ultra-processed foods (UPFs) are increasingly recognized as major contributors to obesity, cardiometabolic disease, and premature death.^4,5^ Together, excessive alcohol use and poor dietary quality represent two of the most important modifiable behavioral determinants of health, and understanding whether these conditions share common mechanisms and frequently co-occur may provide important insights for prevention and treatment of two major public health challenges.

Chronic alcohol exposure and AUD are characterized by alterations in neurocircuitry involved in reward processing, resulting in reduced sensitivity to natural rewards and increased motivation to seek alcohol despite adverse consequences.^6,7^ Similar disruptions in reward processing, inhibitory control, and motivational regulation have also been implicated in obesity, suggesting shared neurobiological mechanisms across these two chronic diseases.^8^ This overlap has prompted growing interest in whether similar addiction-related mechanisms contribute to compulsive eating behaviors, including food addiction (FA), a phenotype characterized by loss of control over food intake, persistent craving, and continued consumption despite adverse consequence.^9,10^ Highly palatable UPFs, particularly those rich in refined carbohydrates and fats, have been proposed to engage reward pathways similar to those implicated in alcohol and other substance use disorders.^11^ Although UPFs have been implicated in addictive-like eating behaviors, only two studies have examined UPF consumption among individuals with AUD, highlighting the limited evidence base in this population.^12,13^ A recent literature review examining associations between UPF consumption, substance use disorders and other addictive behaviors, and addiction recovery outcomes further supports potential overlap between dietary and substance-related addictive processes.^14^

Beyond these shared neurobiological mechanisms, accumulating evidence suggests that AUD and FA are also characterized by overlapping psychological and behavioral vulnerabilities, including depression, anxiety, chronic stress, childhood trauma, emotion regulation difficulties, impulsivity, and sleep disturbance.^15,16^ Together, these shared vulnerabilities provide a strong rationale for expecting common underlying mechanisms across alcohol-related and food-related addictive behaviors. Consistent with this framework, individuals with substance use disorders appear to be particularly vulnerable to food addiction. Previous studies and reviews have reported co-occurrence of addictive eating behaviors and substance use disorders, supporting the possibility that food addiction represents another manifestation of shared addiction-related processes.^17,18^ One study conducted in Italy (the FODRAT study) reported a higher prevalence of food addiction among individuals with alcohol, drug, and tobacco addictions than in the general population.^19^ Outside of AUD populations, food addiction has consistently been associated with greater psychological distress, obesity, and poorer overall health.^20–22^ Determining whether food addiction identifies a similarly vulnerable subgroup among individuals with AUD may have important implications for clinical assessment and treatment.

Additionally, community-based studies further suggests that addictive eating behaviors and alcohol-related problems frequently co-occur, indicating that these behaviors may cluster within vulnerable individuals.^23^ Emerging evidence also suggests that glucagon-like peptide-1 (GLP-1) receptor agonists may reduce both alcohol-related and food-related addictive behaviors, providing additional support for shared biological pathway.^8,24^ The possibility that a single pharmacologic approach may influence both alcohol-related and food-related addictive behaviors further supports the hypothesis that these disorders share overlapping biological mechanisms.

Despite these converging lines of evidence, remarkably little is known about food addiction specifically among individuals diagnosed with AUD. Existing evidence has largely been derived from community samples or mixed substance use disorder populations, leaving the prevalence, clinical characteristics, and psychological correlates of FA within AUD insufficiently characterized. A better understanding of the extent to which food addiction occurs among individuals with AUD may provide insight into shared mechanisms and inform more comprehensive treatment approaches for this high-risk population. The aim of this study was to determine the prevalence of FA symptoms among individuals with AUD and to examine associations between FA symptoms, alcohol-related characteristics, and psychological well-being in populations treatment-seeking and non-treatment-seeking for AUD.

## Methods

### Study Overview

This cross-sectional retrospective analysis examined food addiction (FA) symptoms among adults with AUD enrolled in the National Institute on Alcohol Abuse and Alcoholism (NIAAA) Natural History Protocol (14-AA-0181; ClinicalTrials.gov identifier: NCT02231840). Participants with AUD were enrolled between 2015 and 2025 and included both treatment-seeking (Tx) and non–treatment-seeking (non-Tx) individuals. The study was approved by the Institutional Review Board at the National Institutes of Health (NIH), and all participants provided written informed consent.

#### Participants

Participants were adults meeting Diagnostic and Statistical Manual of Mental Disorders (DSM-IV or DSM-5) criteria for AUD based on structured clinical interviews.^3,25^ Tx participants were admitted to the NIH Clinical Center for inpatient alcohol treatment, whereas non–Tx participants were recruited through community advertisements, evaluated in an outpatient setting, and were not actively seeking or engaged in treatment for AUD at enrollment. All participants completed standardized clinical assessments and self-report questionnaires. Details regarding demographic classifications, smoking status, and questionnaire administration are provided in the **Supplement**. Blood samples were obtained and analyzed for standard clinical chemistry panels, including lipid profiles, glycemic markers, and additional metabolic biomarkers, using routine clinical laboratory assays.

#### Alcohol Use Related Variables

AUD severity was characterized using multiple complementary measures. The number of DSM-IV or DSM-5 AUD (DSM IV from 2015 through 2017; DSM-5 from 2017 through 2025) criteria endorsed was assessed using structured clinical interviews based on the Structured Clinical Interview for DSM-IV (SCID-IV) or the Structured Clinical Interview for DSM-5 (SCID-5), depending on the diagnostic criteria in use at the time of enrollment.^26^ Additional measures of alcohol use severity included total years of heavy drinking as assessed by the Lifetime Drinking History (LDH),^27^ and total scores on the Alcohol Use Disorders Identification Test (AUDIT)^28^ and the Alcohol Dependence Scale (ADS).^29^ Alcohol consumption over the 90 days prior to inpatient admission during outpatient evaluation was quantified using the Timeline FollowBack (TLFB).^30^ Variables derived from the TLFB included average drinks per day, average drinks per drinking day, and number of heavy drinking days.

#### Yale Food Addiction Scale

The Yale Food Addiction Scale (YFAS) is a widely used and well-validated measure of addictive-like eating that was originally developed to operationalize DSM-IV substance dependence criteria in relation to food consumption.^9,31^ The measure has demonstrated strong psychometric properties across diverse populations, including good internal consistency, convergent validity, and clinical utility.^31^ In this report, the term FA refers to food addiction symptoms assessed by the YFAS and does not imply a formally established clinical diagnosis. In this study we classified participants as meeting criteria for FA if they endorsed ≥2 symptoms. This threshold was selected to align the original DSM-IV–based YFAS with current DSM-5 diagnostic conventions for substance use disorders, which require two rather than three symptoms for diagnosis.^3,25^ We did not require endorsement of the clinical impairment/distress criterion because evidence suggests that some individuals who meet symptom thresholds but do not report impairment or distress nevertheless exhibit comparable levels of dysfunction, potentially reflecting limited insight into the impact of their symptoms.^32^ This consideration may be particularly relevant in populations with AUD. Participants endorsing ≥2 YFAS symptoms were therefore classified as having FA in the present analyses.

#### Statistical Analysis

Descriptive statistics were calculated for all study variables and are presented as means and standard deviations (SDs) for continuous variables and frequencies (percentages) for categorical variables.

Differences in demographics, psychological assessments, alcohol-related measures, and smoking status between participants with and without FA were evaluated separately within Tx and non-Tx cohorts using independent-samples t tests for continuous variables and Pearson χ² tests for categorical variables.

Multivariable logistic regression models stratified by treatment seeking status were conducted to examine the associations of alcohol use status and psychological assessments with FA, adjusting for age, education, and income. Adjusted estimates, standard errors, odds ratios (<u>ORs</u>), and 95% confidence intervals (CIs) were reported where applicable. Statistical significance was defined as a 2-sided P < .05. Analyses were conducted using IBM SPSS Statistics, version 29 (IBM Corp., Armonk, NK, USA).

## Results

### Overview of Study Population

The study included 743 individuals with AUD, including 534 Tx and 209 non-Tx participants. Participant characteristics are summarized in **Table S1**. The sample was predominantly male (67.2%), with similar sex distributions across groups. Racial composition differed, with a higher proportion of Black participants in the non-Tx group (52.2%) and White participants in the Tx group (47.4%). The mean (SD) age was 44.1 (12.2) years, with non-Tx participants younger than Tx participants (39.1 [13.1] vs 46.1 [11.3] years, respectively). Body mass index was similar across groups (mean, 27.3 kg/m²). Smoking was more prevalent among Tx participants (59.2% vs 34.0%). Most participants were not married (78.4%), and low income was the most frequently reported category (46.1%). Educational attainment differed, with college graduation more common among non-Tx participants (31.6%). Alcohol use severity was higher in Tx participants, as reflected by greater dependence scores (ADS: 21.68 vs 9.31; AUDIT: 28.54 vs 14.75), longer duration of heavy drinking (14.89 vs 8.35 years), and higher levels of recent alcohol consumption (mean drinks per day: 11.71 vs 4.39; heavy drinking days: 66.05 vs 36.72).

### Food Addiction Symptoms in Adults with Alcohol Use Disorder

Approximately one-third of participants with AUD met criteria for food addiction (YFAS: ≥2) (238 of 743 [32.1%], **Table S1**). The prevalence was similar between non–Tx (64/209, 30.6%) and Tx seeking participants (174/534, 32.6%), (χ²(1) = .27, P = 0.61) (**Figure 1).** Among non-Tx with AUD, those with FA differed significantly from those without with respect to education level (P = 0.02), household income (P = 0.02), and smoking status (P = 0.02), but not sex, race/ethnicity, or marital status (**Table 1**). In contrast, among Tx with AUD, only age (P = .03) and BMI (p <.001) differed between FA, no other demographic variables differed significantly between those with and without FA (**Table 1**). Variables associated with food addiction symptom status in either cohort (age, education, and income) were included as covariates in multivariable logistic regression models.

**Figure 1:**
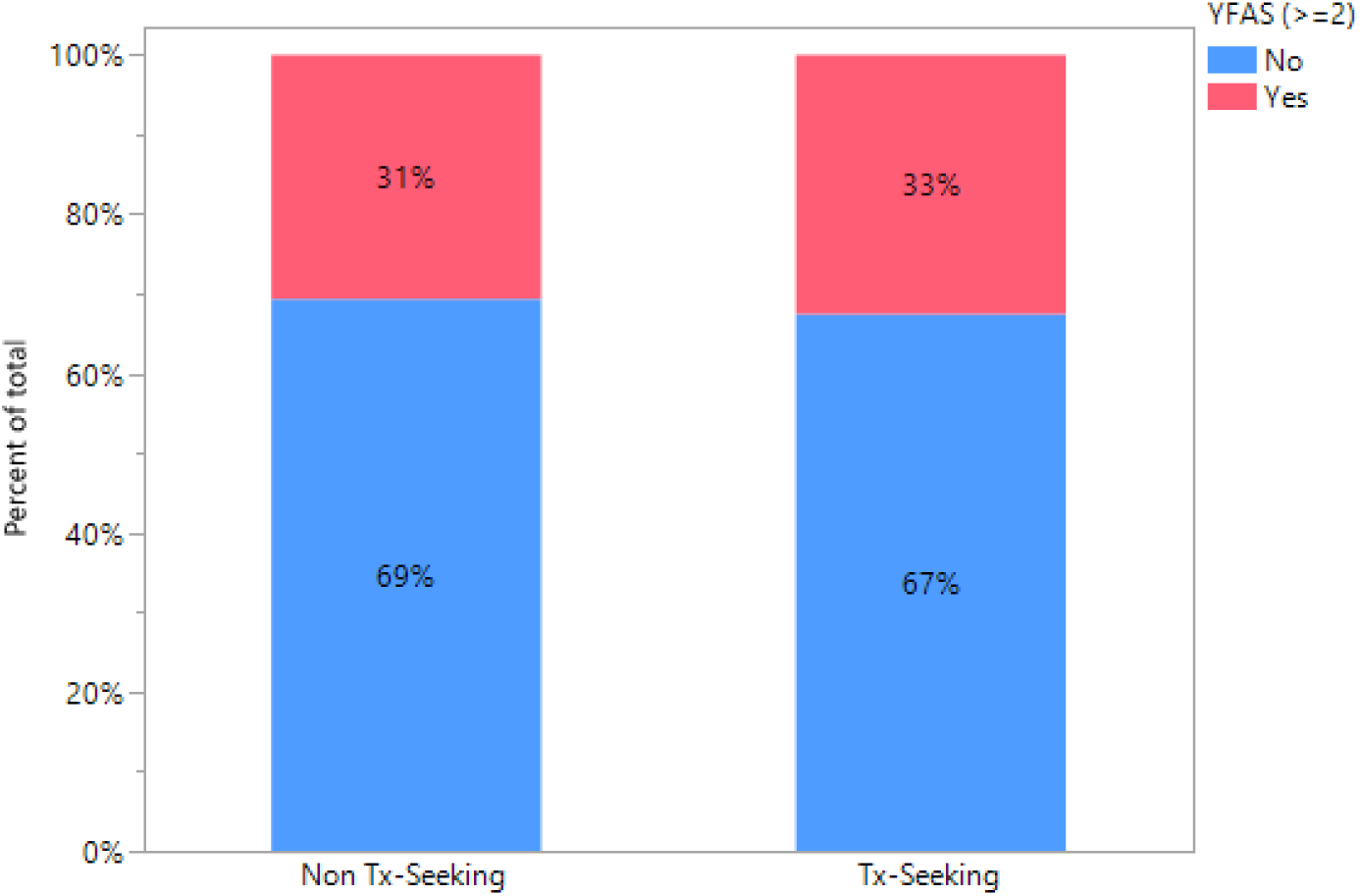
Prevalence of Food Addiction Symptoms Among Non–Treatment-Seeking and Treatment- Seeking Individuals with AUD. Stacked bar chart showing the proportion of participants with food addiction (FA; red) and without food addiction (blue) among treatment-seeking (33%) and non–treatment-seeking (31%) adults with alcohol use disorder. Prevalence difference across the two cohorts is not statistically significant (P = 0.61).

**Table 1:** Demographic Characteristics of Individuals with Alcohol Use Disorder Stratified by Treatment-Seeking Status and Food Addiction Symptoms.

| Characteristic |  | Non-Treatment Seeking |  |  | Treatment Seeking |  |  |
| --- | --- | --- | --- | --- | --- | --- | --- |
|  |  | FA<br>(n=64) | No FA<br>(n=145) | P-value | FA<br>(n=174) | No FA<br>(n=360) | P-value |
| Age, years, mean (SD) |  | 38 (11.98) | 39 (13.55) | .80 | 44.53 (11.43) | 46.79 (11.18) | **.03** |
| BMI, kg/m<sup>2</sup>, mean (SD) |  | 29 (6.24) | 27 (5.19) | **.04** | 28.32 (6.57) | 26.39 (5.33) | **<.001** |
| Sex, n (%) | Female | 19 (29.7) | 50 (34.5) | .50 | 62 (35.6) | 113 (31.4) | .33 |
|  | Male | 45 (70.3) | 95 (65.5) |  | 112 (64.4) | 247 (68.6) |  |
| Race/ethnicity, n (%) | NH-White/Caucasian | 14 (21.9) | 42 (29.0) | .66 | 70 (40.2) | 183 (50.8) | .10 |
|  | NH-Black/African American | 37 (57.8) | 72 (49.7) |  | 73 (42.0) | 115 (31.9) |  |
|  | NH-Other | 8 (12.5) | 17 (11.7) |  | 17 (9.8) | 33 (9.2) |  |
|  | Hispanic | 5 (7.8) | 14 (9.7) |  | 14 (8.0) | 29 (8.1) |  |
| Marital Status, n (%) | Not married | 51 (79.7) | 117 (81.8) | .72 | 136 (79.5) | 274 (76.3) | .41 |
|  | Married/Living as married | 13 (20.3) | 26 (18.2) |  | 35 (20.5) | 85 (23.7) |  |
| Education, n (%) | < HS | 5 (7.8) | 6 (4.2) | **.02** | 20 (11.6) | 37 (10.3) | .32 |
|  | HS graduate | 25 (39.1) | 32 (22.5) |  | 57 (32.9) | 105 (29.3) |  |
|  | Some college/AA | 12 (18.8) | 28 (19.7) |  | 46 (26.6) | 98 (27.4) |  |
|  | College graduate | 18 (28.1) | 47 (33.1) |  | 31 (17.9) | 90 (25.1) |  |
|  | Post college graduate | 4 (6.3) | 29 (20.4) |  | 19 (11.0) | 28 (7.8) |  |
| Household Income, n (%) | Low income | 32 (50.0) | 49 (34.3) | **.02** | 94 (54.0) | 166 (46.4) | .25 |
|  | Medium income | 25 (39.1) | 54 (37.8) |  | 45 (25.9) | 109 (30.4) |  |
|  | High income | 7 (10.9) | 40 (28.0) |  | 35 (20.1) | 83 (23.2) |  |
| Smoking Status (SHQ), n (%) | Smoker | 29 (45.3) | 42 (29.0) | **.02** | 103 (59.2) | 213 (59.2) | >.99 |
|  | Non-smoker | 35 (54.7) | 103 (71.0) |  | 71 (40.8) | 147 (40.8) |  |

Abbreviations: FA=Food addiction; No-FA= no food addiction. Abbreviations: AA=Associate of Arts; BMI=Body Mass Index; HS=High School; NH=Non-Hispanic; SHQ=Smoking History Questionnaire. P -values from students t tests and $\chi^2$ tests. Bold indicates p<.05.

### Clinical Correlates of FA symptoms in Adults with AUD

In unadjusted analyses, individuals with FA in both the non-Tx and Tx cohorts had higher BMI and reported greater childhood trauma, anxiety, perceived stress, depressive symptoms, and poorer sleep quality compared with individuals without FA (**Table 2, Figure S1**). FA was also associated with greater alcohol dependence severity, as measured by the ADS, in both cohorts. In the non-Tx cohort, participants with FA additionally had higher total Alcohol Use Disorders Identification Test (AUDIT) scores than those without FA (mean [SD], 17.27 [7.49] vs. 13.63 [6.39]; *P* < .001), whereas no difference in total AUDIT scores was observed in the Tx cohort. In contrast, AUDIT-C scores were lower among participants with FA in the Tx cohort (mean [SD], 10.16 [2.36] vs 10.64 [1.83]; *P* = .02) but did not differ in the non-Tx cohort. Measures of alcohol consumption, including average drinks per day, drinks per drinking day, years of heavy drinking, and nicotine dependence severity, were not significantly associated with FA in either sample.

**Table 2:** Unadjusted Associations of Clinical and Behavioral Variables with FA in Non-Tx and Tx AUD Cohorts.

| Characteristic | Non-Tx |  |  | Tx |  |  |
| --- | --- | --- | --- | --- | --- | --- |
|  | FA<br>mean (SD) | No FA<br>mean (SD) | P-value | FA<br>mean (SD) | No FA<br>mean (SD) | P-value |
| **Psychological** |  |  |  |  |  |  |
| CTQ | 45.37 (20.10) | 39.58 (16.2) | **.03** | 49.72 (20.47) | 41.2 (15.75) | **<.001** |
| BSA | 4.27 (5.48) | 2.27 (3.27) | **.01** | 14.71 (8.09) | 11.92 (8.01) | **<.001** |
| MADRS | 5.69 (7.53) | 3.01 (4.39) | **.01** | 18.78 (9.59) | 16.46 (10.24) | **.01** |
| PSS | 17.92 (7.32) | 14.14 (6.93) | **<.001** | 23.84 (6.84) | 21.55 (6.86) | **<.001** |
| STAIT | 41.08 (11.14) | 34.49 (9.77) | **<.001** | 51.95 (11.73) | 47.98 (11.22) | **<.001** |
| PSQI | 6.98 (3.33) | 5.56 (3.54) | **.01** | 11.01 (4.44) | 10 (4.05) | **.02** |
| **Alcohol-Related** |  |  |  |  |  |  |
| ADS | 12.06 (7.07) | 8.06 (5.48) | **<.001** | 23.2 (9.04) | 20.96 (8.31) | **.01** |
| AUDIT (Total score) | 17.27 (7.49) | 13.63 (6.39) | **<.001** | 28.39 (7.2) | 28.61 (6.13) | .72 |
| AUDIT (C score) | 7.69 (2.59) | 7.48 (2.21) | .56 | 10.16 (2.36) | 10.64 (1.83) | **.02** |
| 90-day TLFB average drinks per day | 4.97 (4.36) | 4.13 (3.49) | .14 | 11.91 (10.66) | 11.61 (8.72) | .74 |
| 90-day TLFB average drinks per drinking day | 6.8 (4.77) | 6.19 (3.72) | .32 | 15.04 (10.84) | 14.25 (8.47) | .40 |
| 90-day TLFB heavy drinking days | 39.58 (28.74) | 35.46 (28.95) | .34 | 62.66 (28.03) | 67.69 (26.64) | **.05** |
| LDH heavy drinking years | 10.49 (12.34) | 7.37 (11.39) | .09 | 13.74 (11.06) | 15.43 (10.69) | .10 |
| **Smoking Status** |  |  |  |  |  |  |
| Nicotine Dependence (FTND) | 3.55 (2.77) | 2.7 (2.31) | .17 | 3.92 (2.31) | 3.85 (2.44) | .81 |

Note: FA=Food addiction; No FA=no food addiction. Data are presented as mean (SD). Unadjusted *P*-values from independent *t*-tests. ADS=Alcohol Dependence Scale; AUDIT=Alcohol Use Disorder Identification Test; BMI=Body Mass Index; BSA=Brief Scale for Anxiety; CTQ=Childhood Trauma Questionnaire; FTND=Fagerström Test for Nicotine Dependence score; LDH=Lifetime Drinking History; MADRS=Montgomery–Åsberg Depression Rating Scale; Non-Tx=Non-treatment seeking; PSQI=Pittsburgh Sleep Quality Index; STAIT=State-Trait Anxiety, PSS=Perceived Stress Scale; TLFB=Timeline Followback; Tx=Treatment seeking.

In unadjusted analyses of cardiometabolic, hepatic, renal, and glycemic biomarkers (**Table S3**), FA was not associated with most metabolic measures in either cohort. In the Tx group, participants with FA had lower HDL cholesterol (66.71 [27.80] vs 73.14 [31.55] mg/dL; P = .02) and lower estimated glomerular filtration rate using the non–African American equation (94.68 [17.31] vs 99.81 [14.47] mL/min/1.73m²; P = .006). No significant differences were observed for total cholesterol, LDL cholesterol, triglycerides, fasting glucose, HbA1c, blood pressure, heart rate, uric acid, bilirubin, or other kidney function measures in either cohort. In the non-Tx cohort, no biomarker differences were observed between groups.

### Adjusted Associations with FA Symptoms

In both non-Tx and Tx cohorts, several clinical factors were associated with FA symptoms after adjustment for age, education, and income (**Table 3**) (**Figure 2**). Higher BMI, greater anxiety (State-Trait Anxiety Inventory and BSA), higher perceived stress (Perceived Stress Scale), more severe depressive symptoms (MADRS), and greater alcohol dependence severity (ADS) were associated with higher odds of FA symptoms in both cohorts (all P < .05). In adjusted models, only HDL cholesterol measured at Day 2 was significantly associated with FA symptoms in the treatment-seeking cohort (OR, 0.993; 95% CI, 0.986–0.999; P = .026). No other metabolic biomarkers were associated with FA symptoms in either cohort.

**Figure 2:**
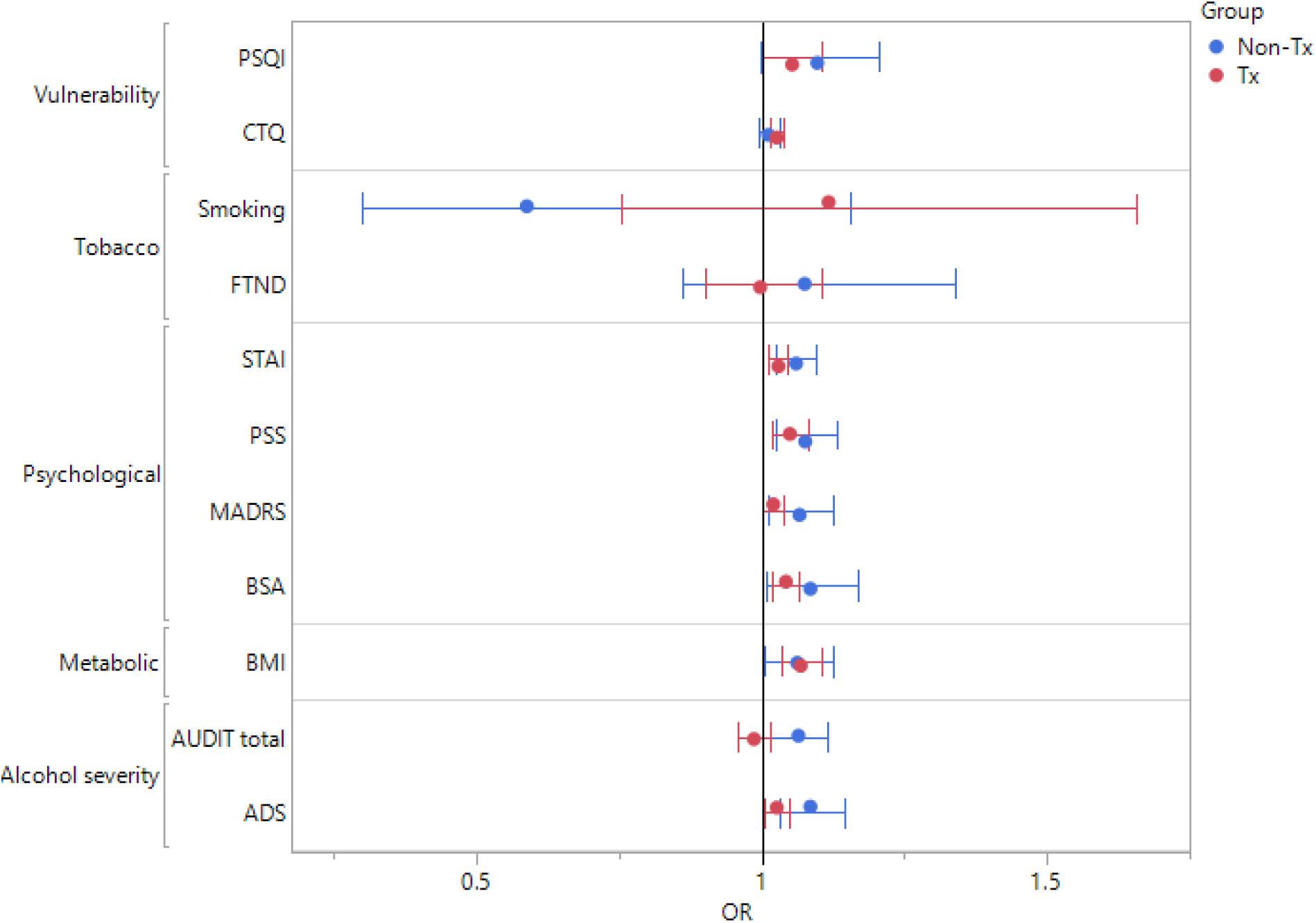
Adjusted Associations Between Clinical Variables and Food Addiction in Individuals with AUD. Forest plot of adjusted odds ratios (ORs) (y-axis) and 95% confidence intervals (CIs) for associations between clinical variables (x-axis) and FA symptoms in treatment-seeking (Tx) (red) and non–treatment-seeking (non-Tx) (blue) individuals with alcohol use disorder (AUD). The vertical reference line denotes no association (OR = 1), with values to the right indicating higher odds of FA symptoms and values to the left indicating lower odds. Models were adjusted for age, education, and income. Points represent ORs, and horizontal lines indicate 95% CIs; the vertical line at OR = 1.0 indicates no association. Variables are grouped by variable domain: trauma, smoking, mental health, health/behavior and alcohol severity. **Abbreviations**: PSQI, Pittsburgh Sleep Quality Index; CTQ, Childhood Trauma Questionnaire; FTND, Fagerström Test for Nicotine Dependence; STAI, State–Trait Anxiety Inventory (trait version); PSS, Perceived Stress Scale; MADRS, Montgomery–Åsberg Depression Rating Scale; BSA, Brief Scale for Anxiety; BMI, body mass index; AUDIT, Alcohol Use Disorders Identification Test; ADS, Alcohol Dependence Scale.

**Table 3:** Adjusted Associations of Clinical and Behavioral Variables with Food Addiction.

| Variable | Non-Treatment Seeking AUD |  |  | Treatment Seeking AUD |  |  |
| --- | --- | --- | --- | --- | --- | --- |
|  | Estimate | SE | *P*-value | Estimate | SE | *P*-value |
| BMI | 0.060 | 0.029 | **.04** | 0.065 | 0.017 | **<.001** |
| **Psychological** |  |  |  |  |  |  |
| CTQ | 0.011 | 0.009 | .24 | 0.026 | 0.005 | **<.001** |
| BSA | 0.082 | 0.038 | **.03** | 0.041 | 0.012 | **<.001** |
| MADRS | 0.064 | 0.028 | **.02** | 0.020 | 0.009 | **.03** |
| PSS | 0.074 | 0.025 | **.003** | 0.048 | 0.015 | **.001** |
| STAIT | 0.058 | 0.017 | **<.001** | 0.028 | 0.009 | **<.001** |
| PSQI | 0.092 | 0.048 | .05 | 0.051 | 0.025 | **.04** |
| **Alcohol-Related** |  |  |  |  |  |  |
| ADS Score | 0.082 | 0.027 | **.002** | 0.025 | 0.012 | **.03** |
| AUDIT total score | 0.062 | 0.024 | **.01** | -0.014 | 0.015 | .35 |
| Audit-C Score | 0.018 | 0.067 | .79 | -0.128 | 0.046 | **.005** |
| 90-day TLFB average drinks per day | 0.025 | 0.044 | .57 | 0.0003 | 0.010 | .98 |
| 90-day TLFB average drinks per drinking day | -0.006 | 0.041 | .89 | 0.006 | 0.010 | .58 |
| 90-day TLFB heavy drinking days | 0.002 | 0.006 | .78 | -0.007 | 0.004 | **.05** |
| LDH heavy drinking years | 0.024 | 0.019 | .20 | -0.010 | 0.011 | .33 |
| **Smoking Status** |  |  |  |  |  |  |
| Smoking (Ref: smoker) | -0.532 | 0.345 | .12 | 0.111 | 0.202 | .58 |
| FTND Score | 0.072 | 0.112 | .52 | -0.003 | 0.053 | .95 |

Note: Multivariable logistic regression models adjusted for age, education, and income. Bold indicates *p*<.05. Abbreviations: ADS, Alcohol Dependence Scale; AUDIT, Alcohol Use Disorders Identification Test; BMI, body mass index; BSA, Brief Scale for Anxiety; CTQ, Childhood Trauma Questionnaire; FTND, Fagerström Test for Nicotine Dependence; LDH, Lifetime Drinking History; MADRS, Montgomery–Åsberg Depression Rating Scale; PSQI, Pittsburgh Sleep Quality Index; PSS, Perceived Stress Scale; STAI-T, State-Trait Anxiety Inventory–Trait; TLFB, Timeline Followback.

Additionally, among the Tx cohort, greater childhood trauma (CTQ) (P <.001) and poorer sleep quality (PSQI) (P = .04) were associated with higher odds of FA symptoms, whereas higher alcohol consumption as measured by the AUDIT-C was inversely associated with FA symptoms (P = .005). In contrast, total AUDIT score was positively associated with FA symptoms only in the non-Tx cohort (P = .01). Smoking status, nicotine dependence severity (Fagerström Test for Nicotine Dependence), average drinks per day, average drinks per drinking day, and years of heavy drinking were not associated with FA symptoms in either cohort (all *P* ≥ .05). (**Table 2).**

## Discussion

In this cross-sectional study of 743 adults with AUD, approximately one-third reported criteria for FA, with similar prevalence observed among treatment-seeking and non–treatment-seeking participants.

To our knowledge, this is the first large-scale study specifically examining FA symptoms in a well-characterized population of adults with AUD. Beside serving as an internal replication, the similar findings across two clinically distinct cohorts suggest that FA occurs across a broad spectrum of AUD rather than being limited to treatment-seeking individuals. Notably, the prevalence observed in this study (32%) was higher than estimates reported in a meta-analysis of community-based populations of approximately 20%.^33^ Although cross-study comparisons should be interpreted cautiously, the higher prevalence in this cohort suggests that individuals with AUD have higher prevalence of FA than broader community-based cohorts. More importantly, FA was consistently associated with greater psychological distress, higher BMI, and greater alcohol dependence severity, suggesting that it may identify a clinically meaningful subgroup of individuals with AUD characterized by greater overall clinical complexity.

Given the substantial health burden associated with both excessive alcohol use and consumption of highly processed, hyperpalatable foods, our results highlight the potential clinical importance of assessing co-occurring addictive eating behaviors in individuals with AUD. While dietary intake behaviors and specifically UPF consumption were not assessed directly in this study, the YFAS assesses addictive eating behaviors toward foods that are often high in sugar and hyperpalatable. Therefore, the present findings should not be interpreted as evidence of dietary intake patterns but rather as evidence that food addiction symptoms are associated with a broader constellation of adverse clinical characteristics. Specifically, individuals with higher food addiction symptoms consistently exhibited greater psychological distress, higher BMI, and greater alcohol dependence severity, suggesting that food addiction may identify a subgroup of individuals with AUD characterized by greater overall clinical complexity and comorbidity.

From a clinical perspective, addictive eating behaviors should not necessarily be viewed as a secondary concern among individuals with AUD but rather as a potential marker of broader behavioral and psychological dysregulation. Routine assessment of FA in both research and clinical settings may improve identification of individuals with greater overall clinical complexity and inform more comprehensive treatment planning. The co-occurrence of AUD and FA also suggests that interventions targeting shared mechanisms in addictive behaviors could provide benefit to both conditions. For example, there is a growing body of evidence supporting a role for GLP-1 therapies in AUD and other addictions (for reviews, see),^24,34,35^ including among individuals with comorbid obesity.^36^ Clinicians should monitor developments in this field as future studies examine whether GLP-1 therapies could provide dual benefit to individuals with co-occurring AUD and FA. The results also support further evaluation of integrated treatment approaches that address alcohol use alongside shared vulnerabilities, including trauma, stress, sleep disturbance, and emotion regulation. Whether targeting these mechanisms improves treatment outcomes remains to be determined.

Across both cohorts, FA was associated with greater psychological distress, higher BMI, and greater alcohol dependence severity. These associations are consistent with evidence that AUD and FA share alterations in reward processing and stress-related pathways.^8,37–39^ Prior research has shown an overlap in the neurobiological systems involved in addiction and a compulsive pattern of food intake, particularly those related to reward processing and motivation.^8,39^ Associations with anxiety, depressive symptoms, and perceived stress were also consistent with stress-vulnerability models of addiction by Sinha et. al, which suggests that chronic stress contributes to both substance use and maladaptive eating behaviors through alterations in reward and self-regulatory processes.^15,16^ The association with higher BMI further highlights the potential cardiometabolic burden of co-occurring FA, although metabolic biomarkers showed little association in the present study.

When cohorts were assessed individually with clinical characteristics, several associations with FA symptoms were identified. Among the Tx group, FA symptoms were additionally associated with greater childhood trauma exposure and poorer sleep quality, findings consistent with prior studies linking both childhood trauma and sleep disturbances to food addiction^40,41^ and with previous work showing a link between childhood trauma exposure and subsequent AUD in adulthood in seeking-treatment individuals.^42^ These findings further support the hypothesis that FA and AUD may arise from shared psychological vulnerabilities rather than representing entirely independent conditions. Alcohol-related measures demonstrated divergent associations according to treatment status. Among treatment-seeking individuals, FA symptoms were inversely associated with AUDIT-C scores, whereas greater alcohol dependence severity was positively associated with FA symptoms among non–treatment-seeking individuals. Indeed, this apparently inconsistent finding may reflect intrinsic differences in motivation to change among individuals with AUD. Specifically, among those not seeking treatment, their addictive behaviors may be less selective and extend across multiple substances or rewards, such as alcohol and food, thereby explaining the positive association between alcohol dependence severity and FA symptoms. In contrast, among treatment-seeking individuals, the inverse association may reflect, at least in part, the use of food as a substitute while attempting to reduce alcohol consumption. Nevertheless, these findings are observational and do not establish causality; thus, these interpretations, while intriguing, remain speculative. Longitudinal studies are needed to clarify these relationships. Future longitudinal studies are needed to determine whether FA influences recovery trajectories, symptom substitution, relapse risk, or treatment response among individuals with AUD.

The YFAS provides a validated framework for assessing addictive eating behaviors using criteria analogous to substance dependence.^9,43^ In the present study, FA clustered with markers of greater clinical severity across both cohorts, suggesting that it may represent a clinically meaningful behavioral phenotype within AUD. Clinically, the observations of this work support routine assessment of addictive eating behaviors as part of a comprehensive evaluation of individuals with AUD. More broadly, the observed overlap between AUD and FA supports investigation of transdiagnostic treatment approaches targeting shared vulnerabilities, including trauma, chronic stress, emotion regulation, and sleep disturbance. In addition, greater integration of nutrition science into AUD treatment, including standardized dietary assessment and evidence-based nutritional interventions, may provide additional benefits for individuals with co-occurring FA symptoms.^44^

This study has several limitations. First, the cross-sectional design precludes conclusions regarding temporal or causal relationships between FA symptoms and clinical characteristics. Second, FA was assessed using the self-reported YFAS rather than a clinical diagnosis. We recognize that the concept and definition of FA remain controversial and are not universally accepted. Although we use the term “food addiction” for simplicity, we do not intend it to imply a formal clinical diagnosis. Rather, throughout this manuscript, FA refers solely to a positive classification on the YFAS, defined as meeting two or more criteria. Third, participants were recruited from a single research protocol, which may limit generalizability to other clinical and community populations. Although analyses were adjusted for key demographic characteristics, residual confounding cannot be excluded.

### Conclusion

Approximately one-third of adults with AUD exhibited FA symptoms regardless of treatment-seeking status. FA was consistently associated with greater psychological distress, higher BMI, and greater alcohol dependence severity, suggesting that it may identify a clinically meaningful subgroup of individuals with AUD characterized by increased overall clinical complexity. Routine assessment of addictive eating behaviors may improve the characterization of AUD in both research and clinical settings and support more comprehensive treatment approaches. This work also highlights potential avenues for the development of integrated treatments for individuals with AUD, particularly those with co-occurring FA symptoms and elevated BMI. For example, in line with a recent meta-analysis,^44^ efforts should be directed toward integrating nutrition science into AUD clinical care, as well as implementing AUD-specific nutritional guidelines and standardized dietary assessment methods. In addition, given the growing body of evidence supporting a role for GLP-1 therapies in AUD and other addictions (for reviews, see ^24,34^), including among individuals with comorbid obesity,^36^ particular attention should be given to their potential dual benefit in people with AUD and FA symptoms. Longitudinal studies are needed to determine potential time and/or causal relationships between AUD and FA, and whether FA influences recovery trajectories, treatment outcomes, and long-term cardiometabolic health.

## Article Information

### Corresponding Authors

Jennifer J. Barb, PhD, MS, Clinical Center, National Institutes of Health, Bethesda, MD, USA; Lorenzo Leggio, MD, PhD, National Institute on Drug Abuse and National Institute on Alcohol Abuse and Alcoholism, National Institutes of Health, Baltimore, MD, USA.

### Authors’ contributions

Drs. Barb, Schwandt and Yang had full access to all the data in the study and take responsibility for the integrity of the data and the accuracy of the data analysis.

*Concept and design*: JJB, LL and AG thought of the basis, rationale, concept, and design.

*Acquisition, analysis, or interpretation of data:* MS, ND and VR were responsible for data acquisition; JJB, LY, MS, LL were responsible for data analysis and interpretation.

*Drafting of the manuscript:* JJB, LY and JY were responsible for original drafting of the manuscript.

*Critical review of the manuscript for important intellectual content:* All authors.

*Statistical analysis:* LY O*btained funding:* LL, ND, JJB

*Administrative, technical, or material support:* JY, LY, JJB.

*Supervision:* JJB, LL.

### Author Disclaimer

This research was supported by the Intramural Research Program of the National Institutes of Health (NIH). The contributions of the NIH authors are considered Works of the United States Government. The findings and conclusions presented in this paper are those of the authors and do not necessarily reflect the views of the NIH or the U.S. Department of Health and Human Services.

### Information on previous presentation of the information reported in the manuscript

This study has not been previously presented up to the date of submission. Preliminary results of this work were presented at the Research Society on Alcohol National Conference in San Antonio, TX on June 22, 2026 and additionally some of the results of this work were featured in the media article in Newswise (https://www.newswise.com/articles/individuals-with-alcohol-use-disorders-often-have-an-unhealthy-relationship-with-their-diet).

### Data Sharing Statement

The data that support the findings of this study are available on request from the corresponding author. However, the data used in this study are drawn from the NIAAA Natural History Protocol, which engages in data sharing through the NIAAA Data Archive (https://nda.nih.gov/niaaa). Access to individual-level data in this repository is restricted, and researchers will need to receive authorization by completing the NDA Data Access Request.

### Additional Contributions

The authors would like to thank the staff at the NIH CC and 1SE unit of the inpatient treatment program for the NIH NIAAA Natural History Protocol. The authors would like to thank Lauren Brick from the NIDA Media team for the figure generation of the graphical abstract.

## References

1. Substance Abuse and Mental Health Services Administration. Key substance use and mental health indicators in the United States: Results from the 2024 National Survey on Drug Use and Health. 2025. HHS Publication No. PEP25-07-007; NSDUH Series H-60. https://www.samhsa.gov/data/data-we-collect/nsduh-national-survey-drug-use-and-health/national-releases

2. Centers for Disease Control and Prevention. Alcohol-Related Disease Impact (ARDI). 2026. https://nccd.cdc.gov/DPH_ARDI/default/default.aspx

3. American Psychiatric Association. Diagnostic and Statistical Manual of Mental Disorders, Fifth Edition (DSM-5). American Psychiatric Association; 2013.

4. Mullen A. Ultra-processed food and chronic disease. Nature Food. 2020/12/01 2020;1(12):771–771. doi:10.1038/s43016-020-00207-3

5. Lane MM, Gamage E, Du S, et al. Ultra-processed food exposure and adverse health outcomes: umbrella review of epidemiological meta-analyses. BMJ. 2024;384:e077310. doi:10.1136/bmj-2023-077310

6. Koob GF, Volkow ND. Neurocircuitry of Addiction. Neuropsychopharmacology. 2010/01/01 2010;35(1):217–238. doi:10.1038/npp.2009.110

7. Volkow Nora D, Koob George F, McLellan AT. Neurobiologic Advances from the Brain Disease Model of Addiction. New England Journal of Medicine. 374(4):363–371. doi:10.1056/NEJMra1511480

8. Leggio L, Farokhnia M, Kenny PJ, Pepino MY, Simmons WK. Crosstalk between alcohol use disorder and obesity: two sides of the same coin? Molecular Psychiatry. 2025/12/01 2025;30(12):5938–5952. doi:10.1038/s41380-025-03259-8

9. Gearhardt AN, Corbin WR, Brownell KD. Preliminary validation of the Yale Food Addiction Scale. Appetite. 2009/04 2009;52(2):430–436. doi:10.1016/j.appet.2008.12.003

10. Gearhardt AN, Schulte EM. Is Food Addictive? A Review of the Science. (1545-4312 (Electronic))

11. Schulte EM, Avena NM, Gearhardt AN. Which foods may be addictive? The roles of processing, fat content, and glycemic load. (1932-6203 (Electronic))

12. Amadieu C, Leclercq S, Coste V, et al. Dietary fiber deficiency as a component of malnutrition associated with psychological alterations in alcohol use disorder. Clin Nutr. May 2021;40(5):2673–2682. doi:10.1016/j.clnu.2021.03.029

13. Barb JJ, King LC, Yang S, et al. An exploratory analysis of the relationship between ultraprocessed food consumption, alcohol intake, body composition, and cardiometabolic markers in individuals with alcohol use disorder. Alcohol Clin Exp Res (Hoboken). Oct 2025;49(10):2184–2198. doi:10.1111/acer.70140

14. Cabral D, Rego M, Freitas-Lemos R, Buckman J. Are Ultra-Processed Foods Associated with Substance Use Behaviors and Addiction Recovery Outcomes? A Narrative Review. Current Addiction Reports. 03/06 2026;13doi:10.1007/s40429-026-00732-4

15. Sinha R, Jastreboff AM. Stress as a Common Risk Factor for Obesity and Addiction. Biological Psychiatry. 2013/05/01/ 2013;73(9):827–835. 10.1016/j.biopsych.2013.01.032

16. Sinha R. Chronic Stress, Drug Use, and Vulnerability to Addiction. Annals of the New York Academy of Sciences. 2008/10/01 2008;1141(1):105–130. 10.1196/annals.1441.030

17. Passeri A, Municchi D, Cavalieri G, Babicola L, Ventura R, Di Segni M. Linking drug and food addiction: an overview of the shared neural circuits and behavioral phenotype. Mini Review. Frontiers in Behavioral Neuroscience. 2023-September-12 2023;Volume 17 - 2023 doi:10.3389/fnbeh.2023.1240748

18. Courbasson CMA, Smith PD, Cleland PA. Substance Use Disorders, Anorexia, Bulimia, and Concurrent Disorders. Canadian Journal of Public Health. 2005/03/01 2005;96(2):102–106. doi:10.1007/BF03403670

19. Tinghino B, Lugoboni F, Amatulli A, et al. The FODRAT study (FOod addiction, DRugs, Alcohol and Tobacco): first data on food addiction prevalence among patients with addiction to drugs, tobacco and alcohol. Eating and Weight Disorders - Studies on Anorexia, Bulimia and Obesity. 03/01 2021;26 doi:10.1007/s40519-020-00865-z

20. Burrows T, Kay-Lambkin F, Pursey K, Skinner J, Dayas C. Food addiction and associations with mental health symptoms: a systematic review with meta-analysis. J Hum Nutr Diet. Aug 2018;31(4):544–572. doi:10.1111/jhn.12532

21. Gearhardt A, Joyner M, Schulte E. Food Addiction. Routl Handbk. 2019:182–191.

22. Gearhardt AN, Schulte EM. Is Food Addictive? A Review of the Science. Annu Rev Nutr. 2021;41:387–410. doi:10.1146/annurev-nutr-110420-111710

23. Hoover LV, Yu HP, Cummings JR, Ferguson SG, Gearhardt AN. Co-occurrence of food addiction, obesity, problematic substance use, and parental history of problematic alcohol use. Psychology of Addictive Behaviors. 2023;37(7):928–935. doi:10.1037/adb0000870

24. Farokhnia M, Leggio L. Prospects of GLP-1 Therapies for Addiction and Mental Health Comorbidities—Quo Vadis?: A Review. JAMA Psychiatry. 03/04 2026;83:306–314. doi:10.1001/jamapsychiatry.2025.4308

25. American Psychiatric Association. Diagnostic and Statistical Manual of Mental Disorders, Fourth Edition, Text Revision (DSM-IV-TR). American Psychiatric Association; 2000.

26. First MB, Williams JBW, Karg RS, Spitzer RL. Structured clinical interview for DSM-5 disorders. Clinician Version (SCID-5-CV). 2015 2015;

27. Skinner HA, Sheu WJ. Reliability of alcohol use indices. The Lifetime Drinking History and the MAST. J Stud Alcohol. 1982/11// 1982;43(11):1157–1170. doi:10.15288/jsa.1982.43.1157

28. Saunders JB, Aasland OG, Babor TF, de la Fuente JR, Grant M. Development of the Alcohol Use Disorders Identification Test (AUDIT): WHO Collaborative Project on Early Detection of Persons with Harmful Alcohol Consumption--II. Addiction. Jun 1993;88(6):791–804. doi:10.1111/j.1360-0443.1993.tb02093.x

29. Skinner HA, Allen BA. Alcohol Dependence Syndrome: Measurement and Validation.

30. Sobell LC, Sobell MB. Timeline follow-back: A technique for assessing self-reported alcohol consumption. Measuring alcohol consumption: Psychosocial and biochemical methods. Springer; 1992:41–72.

31. Meule A, Gearhardt A. Ten Years of the Yale Food Addiction Scale: a Review of Version 2.0. Current Addiction Reports. 09/01 2019;6 doi:10.1007/s40429-019-00261-3

32. Ouellette A-S, Rodrigue C, Lemieux S, Tchernof A, Biertho L, Bégin C. Establishing a food addiction diagnosis using the Yale Food Addiction Scale: A closer look at the clinically significant distress/functional impairment criterion. Appetite. 2018/10/01/ 2018;129:55–61. 10.1016/j.appet.2018.06.031

33. Praxedes DRS, Silva-Júnior AE, Macena ML, et al. Prevalence of food addiction determined by the Yale Food Addiction Scale and associated factors: A systematic review with meta-analysis. Euro Eating Disorders Rev. 2022/03/01 2022;30(2):85-95. 10.1002/erv.2878

34. Srinivasan NM, Farokhnia M, Farinelli LA, Ferrulli A, Leggio L. GLP-1 Therapeutics and Their Emerging Role in Alcohol and Substance Use Disorders: An Endocrinology Primer. Journal of the Endocrine Society. 2025;9(11):bvaf141. doi:10.1210/jendso/bvaf141

35. O’Connor RM. Revisiting food addiction in the era of GLP-1–based obesity pharmacotherapy via neural reward pathways linking feeding and substance use. Review. Frontiers in Behavioral Neuroscience. 2026-April-13 2026;Volume 20 - 2026 doi:10.3389/fnbeh.2026.1805953

36. Klausen MK, Justesen SK, Pedersen JN, et al. Once-weekly semaglutide versus placebo in patients with alcohol use disorder and comorbid obesity: a randomised, double-blind, placebo-controlled trial. The Lancet. 2026;407(10540):1687–1698. doi:10.1016/S0140-6736(26)00305-3

37. Volkow ND, Wang GJ, Tomasi D, Baler RD. Obesity and addiction: neurobiological overlaps. Obesity Reviews. 2013/01/01 2013;14(1):2–18. 10.1111/j.1467-789X.2012.01031.x

38. Kenny PJ. Common cellular and molecular mechanisms in obesity and drug addiction. Nature Reviews Neuroscience. 2011/11/01 2011;12(11):638–651. doi:10.1038/nrn3105

39. Moore CF, Sabino V, Koob GF, Cottone P. Pathological Overeating: Emerging Evidence for a Compulsivity Construct. Neuropsychopharmacology. 2017/06/01 2017;42(7):1375–1389. doi:10.1038/npp.2016.269

40. Hoover LV, Yu HP, Duval ER, Gearhardt AN. Childhood trauma and food addiction: The role of emotion regulation difficulties and gender differences. Appetite. 2022/10/01/ 2022;177:106137. 10.1016/j.appet.2022.106137

41. Li JT, Pursey KM, Duncan MJ, Burrows T. Addictive Eating and Its Relation to Physical Activity and Sleep Behavior. Nutrients. 2018;10(10):1428. doi:10.3390/nu10101428

42. Schwandt ML, Heilig M, Hommer DW, George DT, Ramchandani VA. Childhood Trauma Exposure and Alcohol Dependence Severity in Adulthood: Mediation by Emotional Abuse Severity and Neuroticism. Alcoholism: Clinical and Experimental Research. 2013/06/01 2013;37(6):984–992. 10.1111/acer.12053

43. Meule A, Gearhardt AN. Food Addiction in the Light of DSM-5. Nutrients. 2014;6(9):3653–3671. doi:10.3390/nu6093653

44. Barb JJ, King LC, Nanda S, et al. Dietary intake, quality, and assessment tools in individuals with problematic alcohol use: a scoping review and meta-analysis. Translational Psychiatry. 2026/01/28 2026;16(1):51. doi:10.1038/s41398-026-03842-9

