## Supplemental Materials for "Food Addiction Symptoms in Adults with Alcohol Use Disorder"

**Supplemental Material**

**Supplemental Methods**

***Demographic categorization***
Household income was assessed by self-report and categorized into 3 groups: low income (<$30,000 annually), medium income ($30,000–$74,999), and high income (≥$75,000). Marital status was self-reported as single, married/living as married, divorced, or widowed and recoded as a dichotomous variable: married (married/living as married) and not married (single, divorced, or widowed). Educational attainment was categorized based on years of education as follows: less than high school (0–11 years), high school graduate (12 years), some college or associate’s degree (13–15 years), bachelor’s degree (16 years), and postgraduate or graduate degree (≥17 years). Self-reported race and ethnicity were combined into a single race/ethnicity variable. Participants reporting Hispanic or Latino ethnicity were categorized as Hispanic regardless of race. Among those reporting non-Hispanic ethnicity, participants were classified as non-Hispanic White or non-Hispanic Black based on self-reported race. All other race and ethnicity combinations were categorized as Other.

***Smoking Status Assessments***
Smoking behavior and nicotine dependence were assessed using the Fagerström Test for Nicotine Dependence (FTND) and the Smoking History Questionnaire (SHQ). The FTND is a validated 6-item instrument designed to quantify the intensity of physical nicotine dependence, with total scores ranging from 0 to 10, where higher scores indicate greater dependence.^1^ The SHQ was used to collect detailed self-reported information on smoking behaviors, including age of initiation, duration of smoking, cigarettes smoked per day, and prior quit attempts.^2^ Together, these instruments provided complementary measures of current nicotine dependence and lifetime smoking exposure for all participants included in the analysis.

***Psychological Assessments***

**Perceived Stress Scale (PSS):** Participant perceived stress was assessed using the PSS, a 10-item self-report psychological measure that examines one’s perception of stress over the past 30 days. The PSS is scored on a 5-point Likert scale (0 = never, 1 = almost never, 2 = sometimes, 3 = fairly often, 4 = very often), and PSS total scores can range from 0 to 40, which are then operationalized as low severity (0-13), moderate severity (14-26), and high severity (27-40) of PSS^3^. Average perceived stress over the 30 days prior to admission for inpatient treatment was measured in this study to characterize baseline PSS.

**The Comprehensive Psychopathological Rating Scale (CPRS):** The CPRS was administered at baseline, as well as weekly throughout admission. The CPRS assesses the severity of psychiatric symptoms and observed behaviors with nineteen items that correspond to two subscales: the Brief Scale for Anxiety (BSA) and the Montgomery Asberg Depression Rating Scale (MADRS). After discharge, BSA and MADRS were administered on weeks 1, 2, 4, 8, and 12 as part of an aftercare program for the NIAAA screening protocol. Data was collected for the BSA and MADRS by nurses on the unit from patients who visited the Clinical Center for after care appointments post discharge. The CPRS BSA assesses pathological anxiety alone or combined with other psychological or medical disorders, collected through this10-item measure. The MADRS is a measure that includes 10-items to evaluate the core symptoms of depression. Nine of the ten items on this scale are self-reported, while one is based on the interviewer’s observation during the interview.^4-6^ While not a substitute for clinical diagnoses, MADRS and BSA are used to gauge depression and anxiety symptoms and serve as screening tools.^7,8^

**Childhood Trauma Questionnaire (CTQ):** Childhood trauma exposure was assessed using the validated 25-item CTQ.^9^ The instrument measures emotional abuse, physical abuse, sexual abuse, emotional neglect, and physical neglect occurring before age 18 years. Items are rated on a 5-point Likert scale, generating domain scores (range, 5-25) and a total score (range, 25-125), with higher scores indicating greater trauma exposure. CTQ scores greater than 35 were considered clinically significant.^10^ CTQ scores greater than 35 are typically considered clinically significant with a history of childhood trauma.^11^

**Pittsburgh Sleep Quality Index (PSQI):** The PSQI was used to evaluate sleep quality and disturbance over a 30-day time interval. This 19-item self-administered scale has been validated in normal populations as well as populations with sleep disorders and other psychiatric conditions. The scale sums to a global score on a scale of 0-21. Poor sleep quality is indicated by a global score of 5 or higher, with <5 indicating good sleep quality.^12^

**The State-Trait Anxiety Inventory**: Trait scale is a psychological assessment used to measure trait anxiety, or a person’s tendency to experience anxiety across time and situation. STAI-T is a 20-item measure that quantifies anxiety on a 4-point Likert scale (1 = almost never, 2 = sometimes, 3 = often, 4 = Almost always), and summed scores range from 20-80, where 20-40 typically represents low trait anxiety and 60-80 represents high trait anxiety.

**Supplemental Tables**

**Table S1. Study Population Characteristics**

| **Variable** | | **Total sample (n=743)** | **Non-Tx**  **(n=209)** | **Tx (n=534)** | **Test**  **statistic** | ***P*-value** |
| --- | --- | --- | --- | --- | --- | --- |
| ***Demographics*** | | | | | | |
| Age, years, mean (SD) | | 44.1 (12.2) | 39.13 (13.1) | 46.05 (11.3) | t = -6.7 | **<.001** |
| BMI, kg/m^2^, mean (SD) | | 27.3 (5.8) | 27.85 (5.6) | 27.02 (5.8) | t = 1.8 | .07 |
| Sex, n (%) | Female | 244 (32.8) | 69 (33) | 175 (32.8) | *χ*^2^=0.004 | .95 |
|  | Male | 499 (67.2) | 140 (67) | 359 (67.2) |  |  |
| Race/ethnicity, n (%) | NH-White/Caucasian | 309 (41.6) | 56 (26.8) | 253 (47.4) | *χ*^2^=27.3 | **<.001** |
|  | NH-Black/African American | 297 (40.0) | 109 (52.2) | 188 (35.2) |  |  |
|  | NH-Other | 75 (10.1) | 25 (12.0) | 50 (9.4) |  |  |
|  | Hispanic | 62 (8.3) | 19 (9.1) | 43 (8.1) |  |  |
| Marital status, n (%), n = 737 | Married | 159 (21.6) | 39 (18.8) | 120 (22.6) | *χ*^2^ = 1.3 | .26 |
|  | Not married | 578 (78.4) | 168 (81.2) | 410 (77.4) |  |  |
| Education, n (%), n = 737 | <HS | 68 (9) | 11 (5.3) | 57 (10.7) | *χ*^2^ = 20.2 | **<.001** |
|  | HS graduate | 219 (29.6) | 57 (27.7) | 162 (30.5) |  |  |
|  | Some college/AA | 184 (24.7) | 40 (19.4) | 144 (27.1) |  |  |
|  | College graduate | 186 (25.1) | 65 (31.6) | 121 (22.8) |  |  |
|  | Post college graduate | 80 (0.8) | 33 (16) | 47 (8.9) |  |  |
| Household income, n (%), n = 739 | Low income | 341 (46.1) | 81 (39.1) | 260 (48.9) | *χ*^2^ = 7.1 | **.03** |
|  | Medium income | 233 (31.5) | 79 (38.2) | 154 (28.9) |  |  |
|  | High income | 165 (22.3) | 47 (22.7) | 118 (22.2) |  |  |
| ***Psychological***  mean (SD) | | | | | | |
| CTQ, n = 734 | | 43.2 (17.8) | 41.37 (17.7) | 43.96 (17.6) | t = -1.8 | .08 |
| BSA, n = 740 | | 10.04 (8.5) | 2.88 (4.2) | 12.83 (8.1) | t = -21.8 | **<.001** |
| MADRS, n = 740 | | 13.5 (10.9) | 3.83 (5.7) | 17.15 (10.1) | t = -22.8 | **<.001** |
| PSS, n = 732 | | 20.4 (7.7) | 15.33 (7.3) | 22.29 (6.9) | t = -11.7 | **<.001** |
| STAIT | | 45.7 (12.7) | 36.51 (10.6) | 49.27 (11.5) | t = -14.3 | **<.001** |
| PSQI, n = 633 | | 9.0 (4.5) | 5.99 (3.5) | 10.32 (4.2) | t = -13.4 | **<.001** |
| ***Alcohol-Related***  mean (SD) | | | | | | |
| ADS, n = 734 | | 18.3 (9.8) | 9.31 (6.3) | 21.68 (8.6) | t = -21.4 | **<.001** |
| AUDIT (Total score) | | 24.7 (9.1) | 14.75 (6.9) | 28.54 (6.5) | t = -24.8 | **<.001** |
| AUDIT (C score) | | 9.7 (2.5) | 7.55 (2.3) | 10.48 (2) | t = -16.0 | **<.001** |
| 90-day TLFB average drinks per day | | 9.6 (8.8) | 4.39 (3.8) | 11.71 (9.4) | t = -15.1 | **<.001** |
| 90-day TLFB average drinks per drinking day | | 12.2 (9) | 6.38 (4.1) | 14.50 (9.3) | t = -16.5 | **<.001** |
| 90-day TLFB heavy drinking days | | 57.8 (31) | 36.72 (28.9) | 66.05 (27.2) | t = -12.6 | **<.001** |
| LDH heavy drinking years, n = 704 | | 13.1 (11.4) | 8.35 (11.8) | 14.89 (10.8) | t = -6.7 | **<.001** |
| ***Smoking Status*** | | | | | | |
| SHQ, n (%) | Smoker | 387 (52.1) | 71 (34) | 316 (59.2) | *χ*^2^ = 38.2 | **<.001** |
|  | Non-smoker | 356 (47.9) | 138 (66) | 218 (40.8) |  |  |
| FTND, mean (SD), n = 375 |  | 3.72 (2.4) | 3.06 (2.5) | 3.87 (2.4) | t = -2.53 | .01 |
| ***Yale Food Addiction Scale***  n (%) | | | | | | |
| < 2 Symptom Count (no-FA) | | 505 (68.0) | 145 (69.4) | 360 (67.4) | *χ*^2^ = .27 | .61 |
| ≥ 2 Symptom Count (FA-like) | | 238 (32.0) | 64 (30.6) | 174 (32.6) |  |  |

Note: P values from students t-test and χ² tests. Bold indicates p<.05. Abbreviations: AA=Associate of Arts; ADS=Alcohol Dependance Scale; AUDIT=Alcohol Use Disorder Identification Test; BMI=Body Mass Index; BSA=Brief Scale for Anxiety; CTQ=Childhood Trauma Questionnaire; FTND=Fagerström Test for Nicotine Dependence score; HS=High School; LDH=Lifetime Drinking History; MADRS=Montgomery–Åsberg Depression Rating Scale; NH=Non-Hispanic; Non-Tx=Non-treatment seeking; PSS=Perceived Stress Scale; PSQI=Pittsburgh Sleep Quality Index; SHQ=Smoking History Questionnaire; STAIT=State-Trait Anxiety, TLFB=Timeline Followback, Tx=Treatment seeking.

**Table S2. Unadjusted Associations of Clinical Biomarkers with FA in Non-Tx and Tx AUD Cohorts**

| **Variable** | **Non-Tx** | | | **Tx** | | |
| --- | --- | --- | --- | --- | --- | --- |
|  | **FA**  **mean (SD)** | **No FA**  **mean (SD)** | ***P*-value** | **FA**  **mean (SD)** | **No FA**  **mean (SD)** | ***P*-value** |
| ***Liver Function*** | | | | | | |
| ALT (U/L) | 26.35 (20.55) | 27.66 (26.55) | .73 | 44.67 (48.20) | 58.87 (96.75) | .07 |
| Direct bilirubin (mg/dL) | 0.23 (0.08) | 0.23 (0.08) | .98 | 0.30 (0.28) | 0.33 (0.40) | .51 |
| Total bilirubin (mg/dL) | 0.57 (0.24) | 0.59 (0.28) | .63 | 0.62 (0.50) | 0.66 (0.64) | .54 |
| ***Cardiovascular Measures*** | | | | | | |
| Systolic blood pressure (mmHg) | 123.56 (15.55) | 126.59 (14.56) | .18 | 131.97 (18.86) | 132.51 (16.00) | .73 |
| Diastolic blood pressure (mmHg) | 76.25 (11.60) | 77.16 (10.46) | .58 | 85.91 (16.68) | 85.89 (11.86) | .99 |
| Heart rate (beats/min) | 72.36 (12.33) | 71.77 (12.21) | .75 | 85.55 (17.19) | 87.74 (16.85) | .16 |
| ***Kidney Function*** | | | | | | |
| eGFR-Creatinine (mL/min/1.73m²) | 96.75 (12.31) | 101.50 (18.32) | .66 | 97.67 (16.52) | 100.49 (15.69) | .35 |
| eGFR-Non-AA (mL/min/1.73m²) | 92.44 (16.83) | 95.22 (14.94) | .28 | 94.68 (17.31) | 99.81 (14.47) | **.01** |
| Uric acid (mg/dL) | 5.69 (1.55) | 5.87 (1.66) | .47 | 5.99 (1.74) | 5.91 (1.58) | .57 |
| ***Glucose Regulation*** | | | | | | |
| Fasting glucose (mg/dL) | 94.92 (12.46) | 95.28 (22.69) | .91 | 104.03 (28.01) | 105.48 (28.98) | .58 |
| HbA1c (%) | 5.27 (0.52) | 5.30 (0.81) | .85 | 5.28 (0.50) | 5.32 (0.66) | .56 |
| ***Lipid Profile*** | | | | | | |
| Total cholesterol (mg/dL) | 181.16 (35.43) | 186.92 (39.81) | .32 | 183.64 (44.80) | 185.37 (42.88) | .67 |
| HDL cholesterol (mg/dL) | 62.48 (19.22) | 65.87 (19.35) | .25 | 66.71 (27.80) | 73.14 (31.55) | **.02** |
| LDL cholesterol (mg/dL) | 99.29 (35.28) | 100.33 (35.76) | .85 | 97.02 (38.78) | 93.18 (36.74) | .27 |
| Triglycerides (mg/dL) | 104.06 (90.97) | 104.72 (65.30) | .95 | 110.92 (72.45) | 101.44 (61.48) | .12 |

Note: FA=Food addiction; No FA=no food addiction. Data are presented as mean (SD). Unadjusted *P*-values from independent t-tests. AA=African American; ALT=Alanine aminotransferase; eGFR=Estimated Glomerular Filtration Rate; HbA1c=Hemoglobin A1c; HDL=High-density lipoprotein; LDL=Low-density lipoprotein; Non-Tx=Non-treatment seeking; Tx=Treatment seeking.

**Supplemental Figure**

**Figure S1: Unadjusted Effect Sizes for Clinical and Behavioral Variables According to FA-like Symptom Status in Treatment-Seeking and Non–Treatment-Seeking Adults With Alcohol Use Disorder**

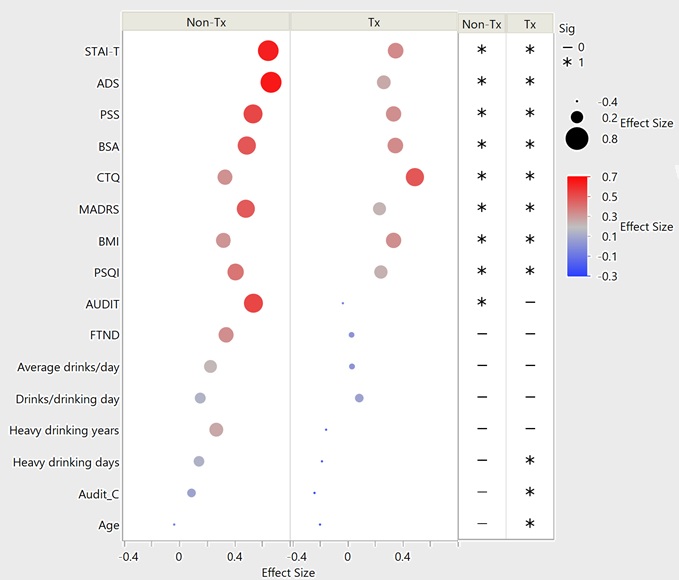

Figure Legend: Dot plot showing standardized effect sizes for differences between participants with and without FA-like symptoms within treatment-seeking (Tx) and non–treatment-seeking (non-Tx) alcohol use disorder cohorts. Bubble size corresponds to the magnitude of the effect size, and color indicates direction of association, with positive values shown in red and negative values shown in blue. Asterisks indicate statistically significant group differences (P < .05). Clinical variables are ordered by effect size magnitude and include measures of anxiety (STAIT), alcohol dependence (ADS), perceived stress (PSS), body shape concerns (BSA), childhood trauma (CTQ), depressive symptoms (MADRS), body mass index (BMI), sleep quality (PSQI), alcohol use severity, and smoking-related variables.

**References**

1.	Heatherton TF, Kozlowski LT, Frecker RC, Fagerstrom K-O. The Fagerström test for nicotine dependence: a revision of the Fagerstrom Tolerance Questionnaire. *British journal of addiction*. 1991 1991;86(9):1119-1127.

2.	Brown RA, Lejuez CW, Kahler CW, Strong DR. Distress tolerance and duration of past smoking cessation attempts. *Journal of Abnormal Psychology*. 2002;111(1):180-185. doi:10.1037/0021-843X.111.1.180

3.	Cohen S, Kamarck T, Mermelstein R. A global measure of perceived stress. *J Health Soc Behav*. Dec 1983;24(4):385-96.

4.	Tyrer PJ, Owen RT, Cicchetti DV. The brief scale for anxiety: A subdivision of the comprehensive psychopathological rating scale. *Journal of Neurology, Neurosurgery & Psychiatry*. 1984;47(9):970-975. doi:10.1136/jnnp.47.9.970

5.	Åsberg M, Montgomery SA, Perris C, Schalling D, Sedvall G. A comprehensive psychopathological rating scale. *Acta Psychiatr Scand*. 1978/04// 1978;57(S271):5-27. doi:10.1111/j.1600-0447.1978.tb02357.x

6.	Montgomery SA, Åsberg M. A New Depression Scale Designed to be Sensitive to Change. *British Journal of Psychiatry*. 1979;134(4):382-389. doi:10.1192/bjp.134.4.382

7.	Hobden B, Schwandt ML, Carey M, et al. The Validity of the Montgomery–Asberg Depression Rating Scale in an Inpatient Sample with Alcohol Dependence. *Alcoholism: Clinical and Experimental Research*. 2017/06/01 2017;41(6):1220-1227. doi:<https://doi.org/10.1111/acer.13400>

8.	Shah NN, Schwandt ML, Hobden B, et al. The validity of the state–trait anxiety inventory and the brief scale for anxiety in an inpatient sample with alcohol use disorder. *Addiction*. 2021/11/01 2021;116(11):3055-3068. doi:<https://doi.org/10.1111/add.15516>

9.	Bernstein DP, Fink L, Handelsman L, et al. Initial reliability and validity of a new retrospective measure of child abuse and neglect. *The American Journal of Psychiatry*. 1994;151(8):1132-1136. doi:10.1176/ajp.151.8.1132

10.	Bernstein D, Stein J, Azzahra F, et al. Development and Validation of a brief screening version of the Chidhood Trauma Questionnaire. *Child abuse & neglect*. 03/01 2003;27:169-90. doi:10.1016/S0145-2134(02)00541-0

11.	Vahapoglu A, Nacar S, Dalgic Y, Gungor H. Is childhood trauma a predictive factor for increased preoperative anxiety levels? *Medicine Science | International Medical Journal*. 01/01 2018:1. doi:10.5455/medscience.2018.07.8928

12.	Buysse DJ, Reynolds CF, Monk TH, Berman SR, Kupfer DJ. The Pittsburgh sleep quality index: A new instrument for psychiatric practice and research. *Psychiatry Research*. 1989/05// 1989;28(2):193-213. doi:10.1016/0165-1781(89)90047-4
